# Symptoms suggestive of COVID-19 in households with and without children: a descriptive survey

**DOI:** 10.1101/2020.11.09.20228205

**Authors:** Grace Grove, Nida Ziauddeen, Nisreen A Alwan

## Abstract

**Background:** Exploring transmission and symptoms of COVID-19 in children is vital, given that schools have recently fully reopened.

**Objectives:** This study aimed to characterise the nature and duration of symptoms suggestive of COVID-19 in UK households, and examine whether the symptoms varied between households with and without children and between adults and children from March to May 2020 in the UK.

**Methods:** An online questionnaire posted on social media (Mumsnet, Twitter, Facebook) was used to gather demographic and symptom information within UK households.

**Results:** Results from 508 households (1057 adults and 398 children) were available for analysis. 64.1% of respondent households with children and 59.1% of households without children had adults with symptoms suggestive of COVID-19. The proportion of adults that reported being symptomatic was 46.1% in households with children (and 36.7% in households without children. In 37.8% of households with at least one adult and one child with symptoms, the child’s onset of symptoms started before the adult. Of all children, 35.7% experienced symptoms, with almost a quarter experiencing fluctuating symptoms for more than 2 weeks compared to almost half of symptomatic adults. In general, children had a shorter (median 5 days) and milder illness course than adults (median 10 days). Fatigue was the most common symptom in adults (79.7%) and cough was the most common symptom in children (53.5%). Chest tightness, shortness of breath, fatigue, muscle ache and diarrhoea were more common in adults than children, while cough and fever were equally common.

**Conclusion:** Children had shorter and milder illness than adults, but in almost a quarter of children symptoms lasted more than 2 weeks. In over a third of both adult-child symptomatic households, the child was the first to become ill. Child to adult transmission and clinical presentation in children need to be further characterised.

**Synopsis:**

- Study question. What is the nature and duration of symptoms suggestive of COVID-19 in UK households with and without children during March and May 2020? Do the symptoms vary between adults and children?
- What’s already known. There has been uncertainty about the extent to which children get and transmit SARS-CoV-2 within households. Symptoms associated with SARS-CoV-2 infection are well described in adults but symptoms and their duration are less well-characterised in children.
- What this study adds On average, children had shorter and milder illness than adults, but still symptoms lasted more than 2 weeks in a significant proportion of children. In over a third of both adult and child symptomatic households, the child was the first to become ill.

## Introduction

Eight months on from the start of the pandemic, there is still ongoing debate around how COVID-19 manifests in children and whether they transmit the infection more or less efficiently than adults. This is particularly important now that schools across the world are have reopened at full capacity. Some suggest that school closures during spring 2020 may have had a modest effect on preventing deaths from COVID-19, much less than other social distancing measures.^1^ However, evidence that children do transmit the virus to each other and to adults is emerging.^2^

A recent COVID-19 outbreak of 260 people in Georgia, USA included 180 children aged 11-17 years and 51 aged 6-10 years who tested positive, with attack rates of 44% and 51% respectively.^3^ A large study of almost 60,000 contacts and 5,700 index cases in South Korea in the period between January and March 2020 found that 17% of contacts have COVID-19 when the index case in the household was aged 10-19 years, compared to an average of 12% for cases of all ages.^4^ A study of 192 children presenting to hospital or urgent care in Massachusetts found nasopharyngeal viral loads in the first days of displaying symptoms to be higher than adults with severe disease.^5^

There is now a wide range of symptoms associated with SARS-CoV-2 infection.^6^ COVID-19 symptoms in children are less well-characterised than in adults. In general, children appear to be more likely to have a mild course than adults.^7,8^ In a recent review, up to 35% of children were asymptomatic. Amongst those who did develop symptoms, the most common was cough, followed by fever and pharyngitis. Other symptoms reported in children include; diarrhoea and vomiting, headache, upper respiratory tract symptoms, wheeze, fatigue, skin signs and less frequently, limb and chest pains.^9,10^ In the Massachusetts study, children with positive and negative SARS-CoV-2 tests had similar symptoms with fever, cough, rhinorrhoea and headache being the most common.^5^ Children can present with severe illness, *Paediatric inflammatory multisystem syndrome associated with SARS-CoV-2* (PIMS-TS) and sadly there have been reported deaths in children.^10^

There is increasing evidence that COVID-19 symptoms in cases who did not require hospitalisation can be protracted and fluctuating. A US study that tracked 292 outpatients who tested positive for SARS-CoV-2 found that 35% of symptomatic respondents reported not returning to their usual state of health by 14-21 days after test date, including 26% among those aged 18-34 years. Continuing symptoms were commonly reported (43% for cough, 35% for fatigue and 29% for shortness of breath).^11^ The COVID Symptom Study which uses a smartphone app to collect COVID-19 symptoms from millions of people found that 1 in 10 of those reporting symptoms is sick for more than three weeks.^12^ A survey in the Netherlands of more than 1,600 mostly non-hospitalised people with COVID-19 symptoms, found that symptoms such as fatigue (88%), shortness of breath (75%), chest pressure (45%), headache (40%), muscle pain (36%) and palpitations (32%) last for months after initial infection.^13^

This study aimed to characterise the nature and duration of symptoms suggestive of COVID-19 in households experiencing mild symptoms, if the symptoms varied between households with and without children, and between adults and children from March to May 2020 in the UK.

## Methods

This is a cross-sectional study using an online questionnaire. Adults aged 18 years and over resident in the UK were eligible to take part in this study. The questionnaire was developed by the research team and published using iSurvey, the University of Southampton’s online survey generation and research tool for distributing online questionnaires. iSurvey stores all information on the University server, which uses secure encryption and ensures that any data cannot be accessed by third parties. Survey responses were anonymous but participants were asked to give consent to be re-contacted in the future regarding COVID-19 testing status and result. Participants who consented were asked to provide an email address and contact telephone number. Ethical approval was granted by the University of Southampton, Faculty of Medicine Ethics Committee (Reference 56462).

The questionnaire was posted on Mumsnet, a parenting website run by parents, and on social media websites (Twitter and Facebook). These websites were chosen to post the questionnaire because it reaches a large audience. Permission was not required to post the questionnaire; however, it could only be posted in one place on Mumsnet (not-for-profit NFP surveys) and could not be shared elsewhere on the website by the researchers. The post contained a link to the questionnaire and brief information about the study. On opening the link, potential participants were taken to an in-depth information sheet. Participants gave their consent by ticking a box and confirmed as part of the consent that they were aged 18 years and over and resident in the UK. They then proceeded on to the questionnaire. Participants were able to share the questionnaire via other social networking sites and email with other people. The questionnaire was posted for a period of two weeks, from May 13^th^ to 25^th^ 2020.

The questionnaire included questions about the adult responding to the questionnaire as well as other adults and children in their household, all of whom are referred to as ‘respondents’ throughout. Questions included demographic information, activities that involved contact with other people, contact with COVID-19 cases and symptoms suggestive of COVID-19 experienced for each household member and their duration.

Data was downloaded from iSurvey once the survey was taken offline. Raw data were checked and blank data rows (indicating participants who had viewed but not completed the questionnaire) were deleted. Participants were asked to report if they were ill on or after 16^th^ March but a large number reported being ill in early March so all participants that reported experiencing symptoms indicative of COVID-19 from the 1^st^ of March were included in the analysis. Households with children were categorised into early years, primary school aged and secondary school aged based on the age of the oldest child in the household.

### Statistical analysis

Statistical analysis was undertaken using Stata 15.0. Descriptive percentages and summary statistics were generated. Univariable comparisons were carried out using an independent t-test for parametric continuous variables, Mann-Whitney test for non-parametric continuous variables and chi square test for categorical variables.

## Results

Responses from a total of 508 households were available for analysis. Data were available on a total of 1057 adults and 398 children (aged 0-18 years). There were 232 households without children, 234 households with children and a further 42 households that were stated as households with children but did not provide information on the children. These 43 households were excluded from analyses comparing households with and without children. Adults in households with children were younger than those in households without children. 36.6% of households without children had at least one member leaving the house for work during lockdown compared to 42.7% of households with children.

The mean number of children in households who reported individual data on children was 1.73 (SD 0.71). There were more respondents in the south than the north of the UK. There were similar proportions of respondents from the north and the south in households with and without children (Table 1). 64.1% of households with children and 59.1% of households without children reported having at least one adult with symptoms indicative of COVID-19. Among households with children, 153 of 234 households reported at least one individual experiencing symptoms, 90 of them had at least one adult and one child experiencing symptoms. Within these 90 households, an adult was first to experience symptoms in 51.1%, a child in 37.8% and an adult and child on the same day in 11.1% (Table S1). In the instances where a child was the first to exhibit symptoms, the proportion was similar in households with primary and secondary children.

**Table 1:** Household characteristics of the study sample

|  | Households without children |  | Households with children |  |
| --- | --- | --- | --- | --- |
|  | n | % | n | % |
| n of households | 232 | - | 234 | - |
| Total n of adults | 488 | - | 496 | - |
| n of adults in household |  |  |  |  |
| 1 | 63 | 27.2 | 19 | 8.1 |
| 2 | 108 | 46.6 | 178 | 76.1 |
| 3 | 36 | 15.5 | 28 | 12.0 |
| 4 | 24 | 10.3 | 7 | 3.0 |
| 5 | 1 | 0.4 | 2 | 0.9 |
| Total n of children | - | - | 398 | - |
| n of children in household |  |  |  |  |
| 1 | - | - | 94 | 40.2 |
| 2 | - | - | 119 | 50.9 |
| 3 | - | - | 18 | 7.7 |
| 4 | - | - | 3 | 1.3 |
| Households categorised by school group |  |  |  |  |
| Early years | - | - | 25 | 10.7 |
| Primary | - | - | 106 | 45.3 |
| Secondary | - | - | 103 | 44.0 |
| Ethnicity of respondent |  |  |  |  |
| White | 215 | 92.7 | 217 | 92.7 |
| BAME | 13 | 5.6 | 15 | 6.4 |
| Unspecified | 4 | 1.7 | 2 | 0.9 |
| Household ethnicity |  |  |  |  |
| White | 205 | 88.4 | 200 | 85.5 |
| BAME | 6 | 2.6 | 6 | 2.6 |
| White and BAME | 19 | 8.2 | 26 | 11.1 |
| Unspecified | 2 | 0.9 | 2 | 0.9 |
| Leaving for work |  |  |  |  |
| No | 147 | 63.4 | 134 | 57.3 |
| Yes | 85 | 36.6 | 100 | 42.7 |
| 1 person | 57 | 24.6 | 70 | 29.9 |
| 2 people | 27 | 11.6 | 30 | 12.8 |
| 3 people | 1 | 0.4 | - | - |
| Region |  |  |  |  |
| South East | 53 | 22.8 | 47 | 20.1 |
| South West | 28 | 12.1 | 16 | 6.8 |
| London | 31 | 13.4 | 52 | 22.2 |
| East of England | 16 | 6.9 | 22 | 9.4 |
| North East | 9 | 3.9 | 2 | 0.9 |
| North West | 26 | 11.2 | 14 | 6.0 |
| East Midlands | 3 | 1.3 | 16 | 6.8 |
| West Midlands | 22 | 9.5 | 15 | 6.4 |
| Yorkshire & Humber | 17 | 7.3 | 13 | 5.6 |
| Scotland | 13 | 5.6 | 24 | 10.3 |
| Northern Ireland | 3 | 1.3 | 5 | 2.1 |
| Wales | 9 | 3.9 | 5 | 2.1 |
| Unspecified | 2 | 0.9 | 3 | 1.3 |
| n of households with at least one symptomatic adult | 137 | 59.1 | 150 | 64.1 |

When looking at individuals, the mean age of adults was 43.9 years (SD 13.4), and the mean age of children was 9.2 years (SD 4.7). 54.0% of the adult respondents were women and 54.8% of the child respondents were boys. Of the total 1057 adults and 398 children, 41.4% (438) of adults and 35.7% (142) children reported being unwell (Table 2). A higher proportion of children (24.1%) than adults (10.5%) described not leaving the home at all during lockdown. The most common reason for leaving the home was exercise for both adults and children. The vast majority of respondents described themselves as being from a white ethnic background (90.7% of adults and 86.9% of children). Out of all adults, 22.9% were leaving home for work.

**Table 2:** Individual characteristics of the study sample for adults and children (aged <18 years)

| Variables | Adults |  | Children |  |
| --- | --- | --- | --- | --- |
|  | n | % | n | % |
| n | 1057 |  | 398 |  |
| n with symptoms | 438 | 41.5 | 142 | 35.7 |
| Age, years (mean ± SD) | 43.9 ± 13.4 | | 9.2 ± 4.7 | |
| Gender |  |  |  |  |
| Male | 477 | 45.1 | 218 | 54.8 |
| Female | 571 | 54.0 | 178 | 44.7 |
| Other | 3 | 0.3 | - | - |
| Unspecified | 6 | 0.6 | 2 | 0.5 |
| Work status during lockdown |  |  |  |  |
| Not leaving for work | 815 | 77.1 | 395 | 99.2 |
| Leaving for work | 242 | 22.9 | 1 | 0.3 |
| Unspecified | - | - | 2 | 0.5 |
| Leaving home during lockdown |  |  |  |  |
| Not leaving the house | 111 | 10.5 | 96 | 24.1 |
| Shopping for essentials | 706 | 66.8 | 36 | 9.0 |
| Exercise | 749 | 70.9 | 299 | 75.1 |
| Caring responsibilities | 67 | 6.3 | - | - |
| Attending school/ nursery | - | - | 29 | 7.3 |
| Ethnicity |  |  |  |  |
| White | 959 | 90.7 | 346 | 86.9 |
| BAME | 63 | 6.0 | 42 | 10.6 |
| Other/Unspecified | 35 | 3.3 | 10 | 2.5 |
| Smoking status |  |  |  |  |
| Non smoker | 821 | 77.7 | - | - |
| Ex-smoker | 120 | 11.4 | - | - |
| Current smoker | 115 | 10.9 | - | - |
| Unspecified | 1 | 0.1 | - | - |

The proportion of adults in households with children that reported being symptomatic (229 of 496, 46.1%) was higher than adults in households without children (179 of 488, 36.7%) (Table 3). Of those adults who experienced symptoms, more were women (60.9% in households without children and 59.4% in households with children). Shortness of breath and sneezing were more common in adults in households without children compared to those with children, with the rest of symptoms being similar (Table 3). A similar pattern was observed between the two household types after excluding households in which at least one individual was leaving home for work during lockdown (Table S2).

**Table 3:** Individual characteristics of symptomatic adults stratified by households with and without children*

|  | Without children |  | With children |  | p-value** |
| --- | --- | --- | --- | --- | --- |
|  | n | % | n | % |  |
| n | 179/488 | 36.7 | 229/496 | 46.1 |  |
| Age, years (mean ± SD) | 45.4 ± 14.5 | | 42.7 ± 8.4 | | **0.02** |
| Gender |  |  |  |  |  |
| Male | 67 | 37.4 | 92 | 40.2 | 0.25 |
| Female | 109 | 60.9 | 136 | 59.4 |  |
| Other | 2 | 1.1 | - | - |  |
| Unspecified | 1 | 0.6 | 1 | 0.4 |  |
| Ethnicity |  |  |  |  |  |
| White | 156 | 87.2 | 209 | 91.3 | 0.60 |
| BAME | 19 | 10.6 | 18 | 7.9 |  |
| Unspecified | 4 | 2.2 | 2 | 0.9 |  |
| Smoking status |  |  |  |  |  |
| Non-smoker | 141 | 78.8 | 182 | 79.5 | 0.98 |
| Ex-smoker | 22 | 12.3 | 27 | 11.8 |  |
| Current smoker | 16 | 8.9 | 20 | 8.7 |  |
| Leaving home during lockdown |  |  |  |  |  |
| None | 14 | 7.8 | 11 | 4.8 | 0.21 |
| Shopping | 126 | 70.4 | 171 | 74.7 | 0.34 |
| Exercise | 125 | 69.8 | 177 | 77.3 | 0.09 |
| Work | 38 | 21.2 | 56 | 24.5 | 0.45 |
| Caring responsibilities | 15 | 8.4 | 14 | 6.1 | 0.38 |
| Duration of symptoms, days (median, IQR) | 14.0<br>(5.0 to 21.0) |  | 10.0<br>(6.0 to 20.0) |  | 0.85 |
| Nature of symptoms |  |  |  |  |  |
| Constant <1 week | 27 | 15.1 | 46 | 20.1 | 0.17 |
| Constant >1 week | 24 | 13.4 | 26 | 11.4 |  |
| Fluctuating <2 weeks | 50 | 27.9 | 46 | 20.1 |  |
| Fluctuating >2 weeks | 76 | 42.5 | 109 | 47.6 |  |
| Unspecified | 2 | 1.1 | 2 | 0.9 |  |
| Sought medical advice for symptomatic adults |  |  |  |  |  |
| No | 94 | 52.5 | 145 | 63.3 | 0.03 |
| Yes | 85 | 47.5 | 84 | 36.7 |  |
| Type of symptoms |  |  |  |  |  |
| Cough | 107 | 59.8 | 119 | 52.2 | 0.13 |
| Fever | 90 | 50.3 | 107 | 46.7 | 0.48 |
| Chest pain | 50 | 27.9 | 59 | 25.9 | 0.64 |
| Chest tightness | 83 | 46.4 | 103 | 45.2 | 0.81 |
| Shortness of breath | 89 | 49.7 | 91 | 39.9 | 0.05 |
| Fatigue | 143 | 79.9 | 185 | 81.1 | 0.75 |
| Muscle ache | 102 | 57.0 | 127 | 55.7 | 0.80 |
| Loss of appetite | 67 | 37.4 | 85 | 37.3 | 0.98 |
| Diarrhoea | 55 | 30.7 | 55 | 24.1 | 0.14 |
| Abdominal Pain | 42 | 23.5 | 40 | 17.5 | 0.14 |
| Vomiting | 11 | 6.2 | 9 | 4.0 | 0.31 |
| Hoarse Voice | 34 | 19.0 | 28 | 12.3 | 0.06 |
| Sneezing | 47 | 26.3 | 30 | 13.2 | **0.001** |
| Loss of sense of smell | 48 | 26.8 | 58 | 25.4 | 0.75 |
| Loss of sense of taste | 45 | 25.1 | 67 | 29.4 | 0.34 |
| Confusion | 17 | 9.5 | 14 | 6.1 | 0.21 |
| Sore throat | 76 | 42.5 | 118 | 51.8 | 0.06 |
| Other: Headache | 11 | 6.2 | 18 | 7.9 | 0.49 |
| Other: Skin | 11 | 6.2 | 12 | 5.3 | 0.71 |

\*Only includes adults who have reported symptoms within the study period excluding 43
households who have reported having children in the household but the respondent provided no data on children)
\*\*Chi square test for categorical variables, Independent t-test or Mann-Whitney test for continuous variables

Overall, 41.4% of adults and 35.7% of children were reported to have symptoms indicative of COVID-19. Of those symptomatic adults, 22.6% were reported to be leaving for work during lockdown while asymptomatic. Fatigue was the most common symptom in adults (79.7% of symptomatic adults) and cough was the most common symptom in children (53.5% of symptomatic children) (Table 4, Figure 1). Presence of cough and fever was similar between adults (55.9% and 48.6% respectively) and children (53.5% and 51.4% respectively). Abdominal pain and vomiting were also similar between adults (20.8%, 5.0%) and children (18.3%, 6.3%). Chest pain, chest tightness and shortness of breath were all more common in adults (26.5%, 45.7%, 44.1% respectively) than in children (4.2%, 7.0%, 11.3% respectively). Adults were also more likely to report muscle ache, loss of appetite, diarrhoea, and loss of sense of taste or smell than children. After excluding households where at least one adult was leaving for work (adults n=258 and children n=69), similar patterns are observed (Table S3).

**Table 4:** Characteristics of symptomatic adults and children (aged <18 years)

| Variables | Adults |  | Children |  | p- value* |
| --- | --- | --- | --- | --- | --- |
|  | n | % | n | % |  |
| n | 438/1057 | 41.4 | 142/398 | 35.7 |  |
| Age, years (mean ± SD) | 43.6 ± 11.5 | | 8.7 ± 4.5 | | **<0.001** |
| Gender |  |  |  |  |  |
| Male | 166 | 37.9 | 76 | 53.5 | **0.004** |
| Female | 268 | 61.2 | 66 | 46.5 |  |
| Other | 2 | 0.5 | - | - |  |
| Unspecified | 2 | 0.5 | - | - |  |
| Work status during lockdown |  |  |  |  |  |
| Not leaving for work | 339 | 77.4 | - | - |  |
| Leaving for work | 99 | 22.6 | - | - |  |
| Leaving home |  |  |  |  |  |
| None | 28 | 6.4 | 25 | 17.6 | **<0.001** |
| Shopping | 316 | 72.1 | 18 | 12.7 | **<0.001** |
| Exercise | 325 | 74.2 | 110 | 77.5 | 0.44 |
| Caring responsibilities | 30 | 6.8 | - | - | - |
| Attending school/ nursery | - | - | 18 | 12.7 | - |
| Duration of symptoms, days | 10.0 |  | 5.0 |  | **<0.001** |

| (median, IQR) | (6.0-21.0) |  | (2.5-8.5) |  |  |
| --- | --- | --- | --- | --- | --- |
| Nature of Symptoms |  |  |  |  |  |
| Constant <1 week | 75 | 17.1 | 47 | 33.1 |  |
| Constant >1 week | 53 | 12.1 | 38 | 26.8 |  |
| Fluctuating <2 weeks | 108 | 24.7 | 23 | 16.2 | **<0.001** |
| Fluctuating >2 weeks | 197 | 45.0 | 32 | 22.5 |  |
| Unspecified | 5 | 1.1 | 2 | 2 |  |
| Sought medical advice |  |  |  |  |  |
| No | 262 | 59.8 | 108 | 76.1 | **<0.001** |
| Yes | 176 | 40.2 | 30 | 21.1 |  |
| Symptoms |  |  |  |  |  |
| Cough | 245 | 55.9 | 76 | 53.5 | 0.60 |
| Fever | 213 | 48.6 | 73 | 51.4 | 0.57 |
| Chest pain | 116 | 26.5 | 6 | 4.2 | **<0.001** |
| Chest tightness | 200 | 45.7 | 10 | 7.0 | **<0.001** |
| Shortness of breath | 193 | 44.1 | 16 | 11.3 | **<0.001** |
| Fatigue | 349 | 79.7 | 68 | 47.9 | **<0.001** |
| Muscle ache | 243 | 55.5 | 28 | 19.7 | **<0.001** |
| Loss of appetite | 160 | 36.5 | 27 | 19.0 | **<0.001** |
| Diarrhoea | 121 | 27.6 | 21 | 14.8 | **0.002** |
| Abdominal Pain | 91 | 20.8 | 26 | 18.3 | 0.52 |
| Vomiting | 22 | 5.0 | 9 | 6.3 | 0.55 |
| Hoarse Voice | 67 | 15.3 | 7 | 4.9 | **0.001** |
| Sneezing | 87 | 19.9 | 16 | 11.3 | **0.02** |
| Loss of sense of smell | 115 | 26.3 | 11 | 7.8 | **<0.001** |
| Loss of sense of taste | 123 | 28.1 | 14 | 9.9 | **<0.001** |
| Confusion | 35 | 8.0 | 2 | 1.4 | **0.005** |
| Sore throat | 208 | 47.5 | 49 | 34.5 | **0.006** |
| Other: Headache | 32 | 7.3 | 10 | 7.0 | 0.97 |
| Other: Skin | 24 | 5.5 | 11 | 7.7 | 0.25 |

\*Chi square test for categorical variables, Independent t-test or Mann-Whitney test for continuous variables

**Figure 1:**
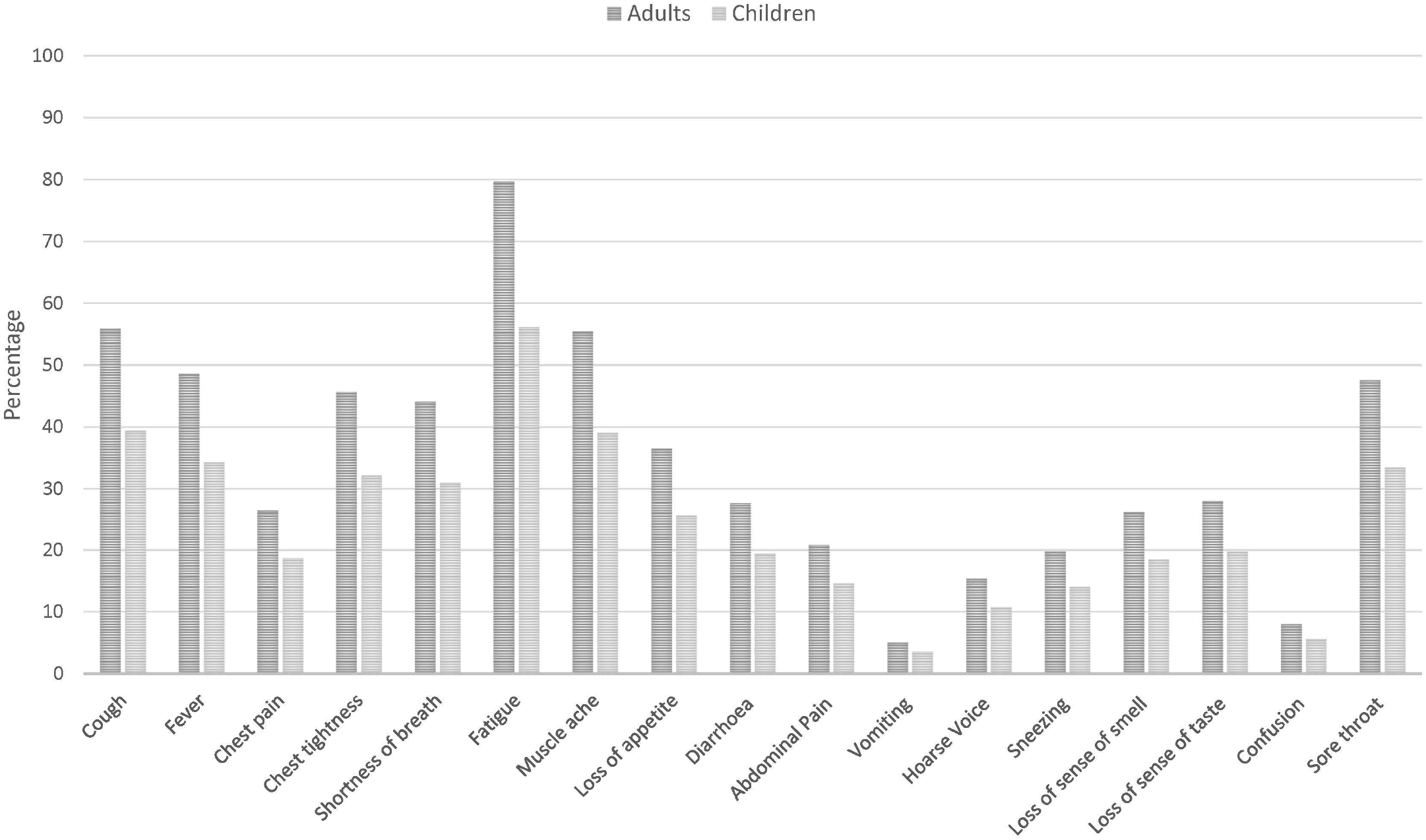
Prevalence of symptoms amongst symptomatic adults (n=438) and children (n=142)

The median duration of symptoms was longer in adults in households without children (14 days, IQR 5.0,21.0) than in adults in households with children (10 days, IQR 6.0, 20.0) (Table 3). 42.5% of the adults in household without children and 47.6% in those with children reported symptoms fluctuating for more than two weeks. Overall, adults (regardless of household status) were unwell for longer (median 10 days, IQR 6.0, 21.0) than children (median 5 days, IQR 2.5, 8.5) (Table 4). Adults were also more likely to report a fluctuating illness for more than two weeks than children (45.0% in adults, 22.5% in children).

Of the adults that reported being tested for SARS-CoV-2, 17 tested positive and 53 tested negative (Table 5). One adult who tested positive reported not experiencing symptoms. Only 7 children were tested, and all received a negative result. Nearly a quarter of those who tested positive were from an ethnic minority background. The majority of individuals who tested positive reported fluctuation in symptoms (31.3% for <2 weeks and 50.0% for >2 weeks, compared to 14.7% and 52.9% respectively in those who tested negative). However, the duration from symptom onset to test was longer in those that tested negative (median 11.5 days, IQR 4, 30) than in those that tested positive (median 4.5 days, IQR 2.5, 6.5). The most common symptoms in those testing positive were cough, fever, fatigue, muscle ache, loss of sense of smell and loss of sense of taste. A high proportion of those who tested negative also reported cough, fever and fatigue as symptoms but muscle ache, loss of sense of smell and loss of sense of taste were less common.

**Table 5:** Characteristics of the adult study sample who were tested for SARS-CoV-2

|  | Tested for SARS-CoV-2 |  |  |  |
| --- | --- | --- | --- | --- |
|  | Tested positive |  | Tested negative |  |
|  | n | % | n | % |
| n | 17 |  | 53 |  |
| Age, years (mean ± SD) | 44 ± 11.0 | | 46.2 ± 12.1 | |
| Gender |  |  |  |  |
| Male | 4 | 23.5 | 20 | 37.7 |
| Female | 13 | 76.5 | 33 | 62.3 |
| Other | - | - | - | - |
| Unspecified | - | - | - | - |
| Ethnicity |  |  |  |  |
| White | 13 | 76.5 | 53 | 100.0 |
| BAME | 4 | 23.5 | - | - |
| Unspecified | - | - | - | - |
| Symptomatic | 16 | 94.1 | 34 | 64.2 |
| Duration from symptom onset to test, median (IQR) | 4.5<br>(2.5 to 6.5) |  | 11.5<br>(4.0 to 30.0) |  |
| Duration of symptoms, median (IQR) | 16.5<br>(9.5 to 27.5) |  | 14<br>(5.0 to 30.0) |  |
| Nature of symptoms |  |  |  |  |
| Constant <1 week | 1 | 6.3 | 3 | 8.8 |
| Constant >1 week | 1 | 6.3 | 8 | 23.5 |
| Fluctuating <2 weeks | 5 | 31.3 | 5 | 14.7 |
| Fluctuating >2 weeks | 8 | 50.0 | 18 | 52.9 |
| Unspecified | 1 | 6.3 | - | - |
| Sought medical advice |  |  |  |  |
| No | 12 | 75.0 | 8 | 23.5 |
| Yes | 4 | 25.0 | 26 | 76.5 |
| Symptoms |  |  |  |  |
| Cough | 11 | 68.8 | 21 | 61.8 |
| Fever | 11 | 68.8 | 23 | 67.6 |
| Chest pain | 5 | 31.3 | 13 | 38.2 |
| Chest tightness | 6 | 37.5 | 19 | 55.9 |
| Shortness of breath | 10 | 62.5 | 19 | 55.9 |
| Fatigue | 16 | 100.0 | 28 | 82.4 |
| Muscle ache | 13 | 81.3 | 16 | 47.1 |
| Loss of appetite | 8 | 50.0 | 10 | 29.4 |
| Diarrhoea | 8 | 50.0 | 9 | 26.5 |
| Abdominal Pain | 3 | 18.8 | 6 | 17.6 |
| Vomiting | - | - | 2 | 5.9 |
| Hoarse Voice | 4 | 25.0 | 9 | 26.5 |
| Sneezing | 5 | 31.3 | 5 | 14.7 |
| Loss of sense of smell | 12 | 75.0 | 4 | 11.8 |
| Loss of sense of taste | 11 | 68.8 | 5 | 14.7 |
| Confusion | 2 | 12.5 | 3 | 8.8 |
| Sore throat | 9 | 56.3 | 16 | 47.1 |
| Other: Headache | 3 | 18.8 | 3 | 8.8 |
| Other: Skin | 1 | 6.3 | 4 | 11.8 |

## Discussion

In this survey, 64.1% of households with children and 59.1% of households without children had adults with symptoms suggestive of COVID-19 in March-April 2020. The proportion of adults that reported being symptomatic was higher in households with children (46.1%) than without children (36.7%). In 37.8% of households with at least one adult and one child with symptoms, the child’s onset of symptoms was before the adult. Of all children, 35.7% experienced symptoms, with almost a quarter of those with symptoms experiencing fluctuating symptoms for more than 2 weeks. In general, children had a shorter and milder illness course than adults. Chest tightness, shortness of breath, fatigue, muscle ache and diarrhoea were more common in adults than children, while cough and fever were equally common.

The main driver for the spread of COVID-19 is person-to-person transmission^14^ but until recently there has been scientific uncertainty around the extent to which children transmit the virus. The evidence that children transmit SARS-CoV-2 is increasing. There was no evidence of difference in rate of infection by age in China^15^ and England.^16^ Household contacts of index cases aged 10-19 years were most likely to test positive (18.6%) followed by household contacts of older adults (17.0% in 60-69 and 18.0% in 70-79 years) and 11.8% overall.^4^ A meta-analysis of nine contact tracing studies concluded that children are half as likely as adults to catch COVID-19. However, the included studies were all considered low to medium quality, included few children, high variation among studies and the adult category included older adults who are known to be at increased risk.^17^

True infection and transmission rate cannot be accurately identified without proactive case finding, mass testing and contact tracing. Children have been tested less frequently as they exhibit milder or no clinical symptoms.^9,18,19^ Two percent of 255,549 identified cases in Italy^20^ and 9.3% of 4,766,825 identified cases in USA^21^ were in children aged 0-18 years. However, the number of child cases in USA showed a 90% increase over 4 weeks from 200,184 on 9^th^ July to 380,174 on 6^th^ August 2020.^21^

Our findings could be supportive of possible viral transmission from children to adults as the symptoms experienced in households were during the weeks around the schools shut down date in England (20^th^ March 2020), when it is likely that community transmission of COVID-19 was reasonably high. In over a third of the surveyed households where at least one adult and one child had symptoms, the child was the first member of the household to display symptoms. Leaving for work during lockdown was more common in households with children, so the index case could have been the person leaving for work in households where there was more than one person unwell, with the child being a secondary case. However, we repeated this analysis excluding the households where adults left the house for work during lockdown as this could be the possible source of infection and the findings were similar. The majority of the clinical research to date has focused on adults, and thus our understanding of the clinical picture of COVID-19 in children is more limited.^7,22^ Additionally, the research that is available on children tends to focus on more severe cases, as these are the group most likely to have been admitted to hospital and tested for COVID-19 early in the pandemic.^23^

Our finding support what has been reported elsewhere that children have shorter and less severe illness than adults.^7,8,22,24,25^ The pathophysiology underlying these differences is unclear, and may include differing immune responses, reduced expression of ACE2 in children (which may be important for viral entry into cells) and increased levels of other viral illnesses in children, leading to competition with SARS-CoV-2^26^. Additionally, children are less likely to have concomitant comorbidities that increase risk of hospitalisation and death from COVID-19^27^, and differences in reporting may impact on symptoms recorded for paediatric cases.

It seems reasonable to assume that young children may find it difficult to clearly vocalise their symptoms, particularly those that are unusual or may be subtle, for example, changes in sense of smell or taste. Interestingly, prevalence of cough and fever was similar between children and adults. Cough and fever are both classic of COVID-19 and clearly detectable symptoms, even for young children. Other studies have also found cough and fever to be common symptoms in children.^22,23,25^ A striking finding in our study has been that almost a quarter of symptomatic children had fluctuating symptoms for longer than two weeks. This survey provided detailed information on symptom type and duration for UK-based households during the time of the COVID-19 pandemic lockdown (which started on 23 March 2020). The household approach allowed exploration of whether adults or children were the first to experience symptoms in their households. However, due to the nature of the online and social media platform this survey may not be representative of the general population, in terms of exposure to SARS-CoV-2 and factors associated with severity of infection such as ethnicity and deprivation.^28^ It is likely those who experienced possible COVID-19 symptoms may have felt more compelled to undertake the survey than those who had not.

One limitation of the household approach was that asking one member of the household to complete the survey for all household members may have resulted in some inaccuracies in responses, if the survey completer relied on their memory rather than asking other household members directly. Additionally, the survey asked for symptom data for those unwell on 16^th^ March or later. However, whilst some participants only gave data for the requested timeframe, many participants provided data for early March (70% in households without children and 75% in households with children reported symptoms on or after 16^th^ March). The decision was made to include these responses in our analysis, but this means that some respondents who were unwell in early March and did not provide data on their symptoms will have been included in the asymptomatic group for the analysis of the data. It was not possible to confirm COVID-19 infection status in most respondents, due to very limited community testing at the time that the survey was undertaken. However, a large number of respondents report both classic COVID-19 symptoms^9,25,29^, and a prolonged, fluctuating course, which is unusual in other infectious illnesses, especially in children. A fluctuating course of respiratory illness in children could be due to co-infection or successive infections. However, this is unlikely to explain the fluctuating course of illness observed in this sample as households were asked to isolate for 14 days and this was followed by lockdown thus restricting exposure to another infection. This prolonged and fluctuating illness, coupled with high community transmission at the time, suggest that it is likely that many of the respondents may truly have had COVID-19. In those who were tested, it is important to consider the timing of test. Those who tested negative were tested later, on average, than those who tested positive. The median time of test for those testing negative was 11.5 days after symptom onset, well beyond the UK government’s recommended 5 days^30^. This may have resulted in some individuals who truly did contract COVID-19 receiving a negative test result as a result of reducing viral load over time.

## Conclusion

Over a third of the children included in this survey experienced symptoms suggestive of COVID-19, and were the first to become unwell in over a third of households where at least one adult and one child were symptomatic. Cough and fever were common symptoms in both adults and children, with almost half of the adults and almost a quarter of the children who became unwell experiencing fluctuating symptoms for more than 2 weeks.

## Data Availability

Anonymised survey data was collected for this study. Ethical approval restricts access to data to the research team. Please contact the research team for further information.

## Acknowledgements

We thank all survey participants for responding to the survey.

