## Supplementary material for "Symptoms suggestive of COVID-19 in households with and without children: a descriptive survey": Table S1

Table S1: First household member to exhibit symptoms within symptomatic households with children

|  | Households with symptoms | Adult | | Child | | | | | Adult and child on same day | | | | |
| --- | --- | --- | --- | --- | --- | --- | --- | --- | --- | --- | --- | --- | --- |
|  |  |  | | All | | Early years | Primary | Secondary | All | | Early years | Primary | Secondary |
|  | n | n | % | n | % | n | n | n | n | % | n | n | n |
| At least one adult or child exhibited symptoms* | 153 | 97 | 63.4 | 46 | 30.1 | 4 | 21 | 21 | 10 | 6.5 | 1 | 3 | 6 |
| March** | 123 | 74 | 60.2 | 41 | 33.3 | 4 | 18 | 19 | 8 | 6.5 | - | 2 | 6 |
| April*** | 27 | 20 | 74.1 | 5 | 18.5 | - | 3 | 2 | 2 | 7.4 | 1 | 1 | - |
| At least one adult and one child exhibited symptoms | 90 | 46 | 51.1 | 34 | 37.8 | 4 | 15 | 15 | 10 | 11.1 | 1 | 3 | 6 |
| March** | 78 | 37 | 47.4 | 33 | 42.3 | 4 | 15 | 14 | 8 | 10.3 | - | 2 | 6 |
| April*** | 12 | 9 | 75.0 | 1 | 8.3 | - | - | 1 | 2 | 16.7 | 1 | 1 | 0 |
| Excluding adults leaving home for work | 43 | 20 | 46.5 | 16 | 37.2 | 1 | 7 | 8 | 7 | 16.3 | 1 | 2 | 4 |
| March** | 40 | 19 | 47.5 | 15 | 37.5 | 1 | 7 | 7 | 6 | 15.0 | - | 2 | 4 |
| April*** | 3 | 1 | 33.3 | 1 | 33.3 | - | - | 1 | 1 | 33.3 | 1 | - | - |
| Also excluding households where children are in school/nursery during lockdown | 34 | 15 | 44.1 | 14 | 41.2 | 1 | 6 | 7 | 5 | 14.7 | - | 1 | 4 |
| March** | 32 | 14 | 43.8 | 13 | 40.6 | 1 | 6 | 6 | 5 | 15.6 | - | 1 | 4 |
| April*** | 2 | 1 | 50.0 | 1 | 50.0 | - | - | 1 | - | - | - | - | - |

*The number first symptomatic in March (123) and April (27) does not add up to the total n (153) as individuals in 3 households were ill in May.

**First symptomatic in March

***First symptomatic in April

Table S2: Individual characteristics of adults stratified by households with and without children* excluding households in which adults are leaving home for work

|  | Without children | | With children | | p-value** |
| --- | --- | --- | --- | --- | --- |
|  | n | % | n | % |  |
|  | 111/294 | 37.8 | 127/301 | 44.2 |  |
| Age, years (mean ± SD) | 47.8 ± 14.6 |  | 42.8 ± 8.2 |  | **0.001** |
| Gender |  |  |  |  |  |
| Male | 39 | 35.1 | 54 | 42.5 | 0.18 |
| Female | 69 | 62.2 | 72 | 56.7 |  |
| Other | 2 | 1.8 | - | - |  |
| Unspecified | 1 | 0.9 | 1 | 0.8 |  |
| Ethnicity |  |  |  |  |  |
| White | 95 | 85.6 | 116 | 91.3 | 0.64 |
| BAME | 13 | 11.7 | 9 | 7.1 |  |
| Unspecified | 3 | 2.7 | 2 | 1.6 |  |
| Smoking status |  |  |  |  |  |
| Non-smoker | 87 | 78.4 | 104 | 81.9 | 0.61 |
| Ex-smoker | 14 | 12.6 | 11 | 8.7 |  |
| Current smoker | 10 | 9.0 | 12 | 9.5 |  |
| Leaving home during lockdown |  |  |  |  |  |
| None | 12 | 10.8 | 9 | 7.1 | 0.31 |
| Shopping | 80 | 72.1 | 96 | 75.6 | 0.54 |
| Exercise | 80 | 72.1 | 105 | 82.7 | 0.05 |
| Caring responsibilities | 8 | 7.2 | 7 | 5.5 | 0.59 |
| Duration of symptoms, days (median, IQR) | 14.0  (6.0 to 21.0) |  | 10.0  (6.0 to 21.0) |  | 0.80 |
| Nature of symptoms |  |  |  |  |  |
| Constant <1 week | 15 | 13.5 | 24 | 18.9 | 0.61 |
| Constant >1 week | 11 | 9.9 | 11 | 8.7 |  |
| Fluctuating <2 weeks | 32 | 28.8 | 30 | 23.6 |  |
| Fluctuating >2 weeks | 51 | 45.9 | 61 | 48.0 |  |
| Unspecified | 2 | 1.8 | 1 | 0.8 |  |
| Sought medical advice for symptomatic adults |  |  |  |  |  |
| No | 56 | 50.4 | 80 | 63.0 | 0.05 |
| Yes | 55 | 49.6 | 47 | 37.0 |  |
| Type of symptoms |  |  |  |  |  |
| Cough | 66 | 59.5 | 71 | 55.9 | 0.58 |
| Fever | 47 | 42.3 | 61 | 48.0 | 0.38 |
| Chest pain | 28 | 25.2 | 28 | 22.1 | 0.56 |
| Chest tightness | 49 | 44.1 | 64 | 50.4 | 0.34 |
| Shortness of breath | 55 | 49.6 | 46 | 36.2 | **0.04** |
| Fatigue | 87 | 78.4 | 103 | 81.1 | 0.60 |
| Muscle ache | 63 | 56.8 | 66 | 52.0 | 0.46 |
| Loss of appetite | 42 | 37.8 | 48 | 37.8 | 1.00 |
| Diarrhoea | 34 | 30.6 | 26 | 20.5 | 0.07 |
| Abdominal Pain | 24 | 21.6 | 20 | 15.8 | 0.24 |
| Vomiting | 8 | 7.2 | 6 | 4.7 | 0.42 |
| Hoarse Voice | 19 | 17.1 | 15 | 11.8 | 0.24 |
| Sneezing | 31 | 27.9 | 15 | 11.8 | **0.002** |
| Loss of sense of smell | 25 | 22.5 | 30 | 23.6 | 0.77 |
| Loss of sense of taste | 26 | 23.4 | 39 | 30.7 | 0.21 |
| Confusion | 7 | 6.3 | 6 | 4.7 | 0.59 |
| Sore throat | 48 | 43.2 | 73 | 57.5 | **0.03** |
| Other: Headache | 6 | 5.4 | 13 | 10.3 | 0.17 |
| Other: Skin | 4 | 3.6 | 9 | 7.1 | 0.23 |

*Only includes adults who have reported symptoms within the study period excluding 43 households who have reported having children in the household but the respondent provided no data on children)

**Chi square test for categorical variables, Independent t-test or Mann-Whitney test for continuous variables

Table S3: Characteristics of symptomatic adults and children (aged <18 years) excluding households in which adults are leaving home for work

| Variables | Adults | | Children | | p-value** |
| --- | --- | --- | --- | --- | --- |
|  | n | % | n | % |  |
| N with symptoms | 258/619 | 41.7 | 69/227 | 30.4 |  |
| Age, years (mean ± SD) | 44.8 ± 11.9 |  | 8.4 ± 4.8 |  | **<0.001** |
| Gender |  |  |  |  |  |
| Male | 97 | 37.6 | 34 | 49.3 |  |
| Female | 157 | 60.9 | 35 | 50.7 | 0.19 |
| Other | 2 | 0.8 | - | - |  |
| Unspecified | 2 | 0.8 | - | - |  |
| Leaving home during lockdown |  |  |  |  |  |
| Not leaving the house | 24 | 9.3 | 12 | 17.4 | 0.06 |
| Shopping for essentials | 188 | 72.9 | 5 | 7.2 | **<0.001** |
| Exercise | 198 | 76.7 | 59 | 85.5 | 0.12 |
| Caring responsibilities | 16 | 6.2 | - | - | - |
| Attending school/ nursery | - | - | 1 | 1.4 | - |
| Duration of symptoms, days (median, IQR) | 12 (6-21) |  | 5 (3-10) |  | **<0.001** |
| Nature of Symptoms |  |  |  |  |  |
| Constant <1 week | 41 | 15.9 | 24 | 34.8 |  |
| Constant >1 week | 24 | 9.3 | 13 | 18.8 | **<0.001** |
| Fluctuating <2 weeks | 71 | 27.5 | 13 | 18.8 |  |
| Fluctuating >2 weeks | 119 | 46.1 | 19 | 27.5 |  |
| Unspecified | 3 | 1.2 | - | - |  |
| Sought medical advice |  |  |  |  |  |
| No | 150 | 58.1 | 53 | 76.8 | **0.001** |
| Yes | 108 | 41.9 | 13 | 18.8 |  |
| Unspecified | - | - | 3 | 4.3 |  |
| Symptoms |  |  |  |  |  |
| Cough | 149 | 57.8 | 42 | 60.9 | 0.80 |
| Fever | 118 | 45.7 | 39 | 56.5 | 0.11 |
| Chest pain | 61 | 23.6 | 2 | 2.9 | **<0.001** |
| Chest tightness | 123 | 47.7 | 4 | 5.8 | **<0.001** |
| Shortness of breath | 112 | 43.4 | 7 | 10.1 | **<0.001** |
| Fatigue | 205 | 79.5 | 33 | 47.8 | **<0.001** |
| Muscle ache | 140 | 54.3 | 10 | 14.5 | **<0.001** |
| Loss of appetite | 95 | 36.8 | 16 | 23.2 | **0.03** |
| Diarrhoea | 69 | 26.7 | 10 | 14.5 | **0.04** |
| Abdominal Pain | 51 | 19.8 | 11 | 15.9 | 0.47 |
| Vomiting | 16 | 6.2 | 6 | 8.7 | 0.46 |
| Hoarse Voice | 38 | 14.7 | 4 | 5.8 | 0.05 |
| Sneezing | 51 | 19.8 | 7 | 10.1 | 0.06 |
| Loss of sense of smell | 59 | 22.9 | 5 | 7.2 | **0.004** |
| Loss of sense of taste | 72 | 27.9 | 5 | 7.2 | **<0.001** |
| Confusion | 16 | 6.2 | 1 | 1.4 | 0.11 |
| Sore throat | 134 | 51.9 | 24 | 34.8 | **0.011** |
| Other: Headache | 21 | 8.1 | 5 | 7.2 | 0.87 |
| Other: Skin | 14 | 5.4 | 3 | 4.3 | 0.83 |

*Chi square test for categorical variables, Independent t-test or Mann-Whitney test for continuous variables
